# Diagnostic accuracy of point-of-care urine tenofovir test, and associations between metrics of tenofovir use and treatment outcomes in a community ART programme

**DOI:** 10.64898/2026.08.13.26360358

**Authors:** Lisanthini Naidu, Kwena Tlhaku, Katya Govender, Yukteshwar Sookrajh, Pravikrishnen Moodley, Johan van der Molen, Natasha Samsunder, Lara Lewis, Monica Gandhi, Paul K. Drain, Christopher C Butler, Gail Hayward, Nigel Garrett, Jienchi Dorward

## Abstract

**Background:** Urine tenofovir (uTFV) and dried blood spot (DBS) tenofovir diphosphate (TFV-DP) concentrations respectively estimate short- and medium-term adherence to tenofovir disoproxil fumarate (TDF)-based antiretroviral therapy (ART). We evaluated the accuracy of a point-of-care uTFV assay, and associations between uTFV/TFV-DP, and viral load (VL) and retention outcomes within a South African community ART programme.

**Methods:** We measured uTFV and DBS TFV-DP concentrations using liquid chromatography-tandem mass spectrometry (LC-MS/MS). We calculated sensitivity and specificity of the point-of-care uTFV assay at the manufacturer-recommended threshold of ≥1,500 ng/mL compared to LC-MS/MS. We assessed associations of the point-of-care uTFV assay, and DBS TFV-DP concentrations with concurrent viraemia, and with retention-in-care by 16 weeks post-enrolment.

**Results:** Of 196 adults median age was 44 years, 127 (64.8%) were female, and 191 (97.4%) were receiving TDF. 185 (94.4%) had detectable point-of-care uTFV, which had high sensitivity (99.5%, 95% CI 96.5-100%) and moderate specificity (76.9%, 95% CI 46.0-93.8%) for detecting uTFV ≥1,500 ng/mL. Two participants had concurrent viraemia ≥1,000 copies/mL; of these 50.0% (95% CI 9.4-90.5) had undetectable point-of-care uTFV, and 100% (95% CI 19.7-100) had low TFV-DP <483 fmol/punch. Among participants without viraemia 97.4% (95% CI 93.6-99.0) had detectable uTFV, and 98.1% (95.3-99.6) had high TFV-DP ≥483 fmol/punch. Point-of-care uTFV and DBS TFV-DP were not associated with retention-in-care.

**Conclusions:** The point-of-care uTFV assay demonstrated high sensitivity and moderate specificity to detect uTFV. Over 95% of people without viraemia had detectable point-of-care uTFV or high DBS TFV-DP levels respectively, but these were not associated with 16-week retention-in-care.

## Introduction

Community antiretroviral therapy (ART) delivery programmes, where ART is collected at community pick-up points instead of clinics, benefit clients and are increasingly common in global HIV treatment programmes [1]. Using point-of-care assays to monitor adherence may improve HIV treatment monitoring in these community programmes [2]. A point-of-care urine tenofovir test (Abbott, Illinois, USA) has been developed. This assay detects urine tenofovir concentrations at a threshold of 1500ng/mL, which has been shown to correspond to whether someone has ingested tenofovir disoproxil fumarate (TDF) in the past 3-4 days [3]. However, there are few studies available regarding the diagnostic accuracy of the point-of-care assay at this threshold, and whether the point-of-care could be used to detect and/or rule out viraemia in community ART programmes. Furthermore, these tests assess short-term adherence, whereas tenofovir diphosphate (TFV-DP) concentrations in dried blood spots (DBS) estimates adherence over the past 6-8 weeks,[4] which may be more strongly associated with outcomes. We aimed to determine the diagnostic accuracy of this point-of-care urine tenofovir assay at the manufacturer’s threshold of 1,500 ng/mL. We also assessed whether urine tenofovir and DBS TFV-DP concentrations were associated with concurrent viraemia, and future retention-in-care at 16 weeks.

## Methods

### Study Design, Setting and Participants

We conducted a prospective diagnostic accuracy sub-study nested within the PHILA point-of-care viral load (VL) clinical trial in South Africa. The PHILA trial was an open-label, individually randomised study evaluating point-of-care VL testing in community ART delivery programmes [5].

Adults with HIV receiving ART through community pick-up points were enrolled at their next clinic visit and followed up for 16 weeks. At enrolment, participants provided sociodemographic and clinical details, including self-reported adherence. Urine, plasma and dried blood spot (DBS) specimens were taken and stored at −80 degrees Celsius.

### Laboratory Procedures

Urine samples were defrosted and tested retrospectively using the Abbott lateral flow point-of-care uTFV assay, according to manufacturer’s instructions at the Africa Health Research Institute (AHRI), Durban, South Africa. Results were read after 3-5 minutes by laboratory staff, with photographs of results reviewed by a clinician, and discrepancies adjudicated by a third investigator. For the reference standard, uTFV was quantified retrospectively using liquid chromatography-tandem mass spectrometry (LC-MS/MS) at AHRI. For TFV-DP, DBS samples were defrosted and TFV-DP concentrations were quantified using LC-MS/MS, also at AHRI (see supplementary material for detailed LC-MS/MS methods).

Plasma HIV-1 VL was measured using the cobas® HIV-1 assay (Roche, Basel, Switzerland) on the cobas 6800 platform.

Staff performing and reviewing LC-MS/MS, VL testing and point-of-care uTFV results were blinded to other results, respectively.

### Diagnostic accuracy variables, outcomes and exposures

For the diagnostic accuracy assessment, the index test was the point-of-care uTFV assay, and the reference standard was uTFV below or above the manufacturer threshold of 1500ng/mL, measured by LC-MS/MS. To assess associations between TFV metrics and clinical outcomes, we defined the outcomes of concurrent viraemia as ≥1,000 copies/mL (primary) and ≥50 copies/mL (secondary) and retention-in-care as ART collection from community pick-up points or in the clinic, by 16 weeks post-enrolment. Exposures were point-of-care uTFV results, and DBS TFV-DP results. For TFV-DP, we used thresholds of 483 and 686 fmol/punch, which we previously found to accurately predict viraemia at 1000 copies/mL and 50 copies/mL, respectively [6].

### Statistical Analysis

To assess diagnostic accuracy of the point-of-care uTFV assay we calculated sensitivity, specificity, positive predictive value (PPV), and negative predictive value (NPV), each with 95% Wilson’s confidence intervals (CIs) with Yates continuity correction. For discrepant results, LC-MS/MS concentrations were reported to explore misclassification around the manufacturer’s threshold of 1,500 ng/mL.

To assess associations between the exposures of point-of-care uTFV results, and DBS TFV-DP concentrations with the outcome of concurrent viraemia, we constructed 2×2 tables and estimated conditional proportions with 95% CIs. To assess associations between point-of-care uTFV results and retention-in-care, we constructed 2×2 tables with Haldane-Anscombe correction (adding 0.5 to each cell) to account for zero-count cells for calculation of odds ratios (ORs) [7] and the delta method for 95% CIs. For DBS TFV-DP concentrations and retention-in-care, we used a logistic regression model with TFV-DP as a continuous variable. People with missing VL or point-of-care results were excluded from respective analyses.

In post-hoc analyses to evaluate the association between point-of-care uTFV results and TFV-DP, we summarised TFV-DP by detectable vs. not detectable uTFV results and compared median TFV-DP between groups using the Mann-Whitney U test.

Sample size was determined by the number of participants enrolled in the PHILA trial who had a point-of-care uTFV result or were receiving TDF-based ART. Analyses were conducted using R version 4.2.0 (R Foundation for Statistical Computing, Vienna, Austria).

### Ethical Considerations

The PHILA trial and this sub-analysis was approved by the University of KwaZulu-Natal Biomedical Research Ethics Committee (BREC/00000837/2019) and the University of Oxford Tropical Research Ethics Committee (OxTREC 64-19). Written informed consent was obtained from all participants prior to enrolment. PHILA is registered with the Pan African Clinical Trials Registry (PACTR202002785960123).

## Results

### Baseline characteristics

Between 15 August 2022 and 24 August 2023, 200 people were enrolled into PHILA, of whom 196 had a point-of-care uTFV result for inclusion in our analysis (Figure S1). Median (IQR) age was 44 years (38.0 to 49.0), 127 were female (64.8%) and median time on ART was 8 years (6 to 10.9). Of these, 191 (97.4%) were taking TDF-based ART and 5 (2.6%) other regimens (Table S1).

### Diagnostic accuracy of the point-of-care uTFV test

The proportion who had a point-of-care uTFV result of detected was 94.4% (185/196), while 93.4% (183/196) had reference standard LC-MS/MS uTFV concentrations ≥1500 ng/mL. The point-of-care uTFV test had high sensitivity (99.5%, 95% CI 96.5-100%) and moderate specificity (76.9%, 95% CI 46.0-93.8%) for detecting uTFV (Table 1). Discrepant results had LCMS uTFV levels between 641-2040 ng/mL (Table S2), with post-hoc review of result pictures showing potential faint lines on the lateral flow assay that may have been missed (Figure S2).

**TABLE 1:** ANALYTIC PERFORMANCE OF THE POINT-OF-CARE URINE TENOFOVIR TEST TO DETECT URINE TENOFOVIR CONCENTRATIONS AT THE MANUFACTURER THRESHOLD OF 1500NG/ML.

|  |  | LC-MS/MS urine tenofovir (uTFV) (ng/mL) |  |  |
| --- | --- | --- | --- | --- |
|  |  | <1500 | ≥1500 | Total |
| **POC TFV** | uTFV not detected | 10 | 1 | 11 |
|  | uTFV detected | 3 | 182 | 185 |
|  | *Total* | 13 | 183 | 196 |
| Sensitivity |  | 99.45% (96.53-99.97) |  |  |
| Specificity |  | 76.92% (45.98-93.84) |  |  |
| PPV |  | 98.38% (94.95-99.58) |  |  |
| NPV |  | 90.91% (57.12-99.52) |  |  |

LC-MS/MS = liquid chromatography tandem mass spectrometry, TFV = tenofovir, POC = point-of-care, PPV = positive predictive value, NPV = negative predictive value

### Associations with concurrent viraemia and future retention-in-care

Of the 191 adults who were on TDF-containing ART, the proportion who had an LC-MS/MS DBS TFV-DP above 483 fmol/ punch was 97.4% (186/191) and above 686 fmol/ punch was 88% (168/191). Regarding viraemia, the proportion who had viraemia at ≥1000 copies/mL was 1.05% (2/191) and >50 copies was 47.1% (9/191). The proportion retained-in-care was 88% (169/191).

Regarding concurrent viraemia, the analysis is limited by the low prevalence (1.0%, 2/191) of viraemia ≥1,000 copies/mL. Point-of-care uTFV was undetectable in 50.0% (95% CI 9.4-90.5) of those with viraemia, and detectable in 97.4% (95% CI 93.6-99.0) of those without viraemia. TFV-DP was <483 fmol/punch for 100% (95% CI 19.7-100) of those with viraemia, and ≥483 fmol/punch for 98.1% (95% CI 95.3-99.6) of those without viraemia (Table 2).

**TABLE 2:**
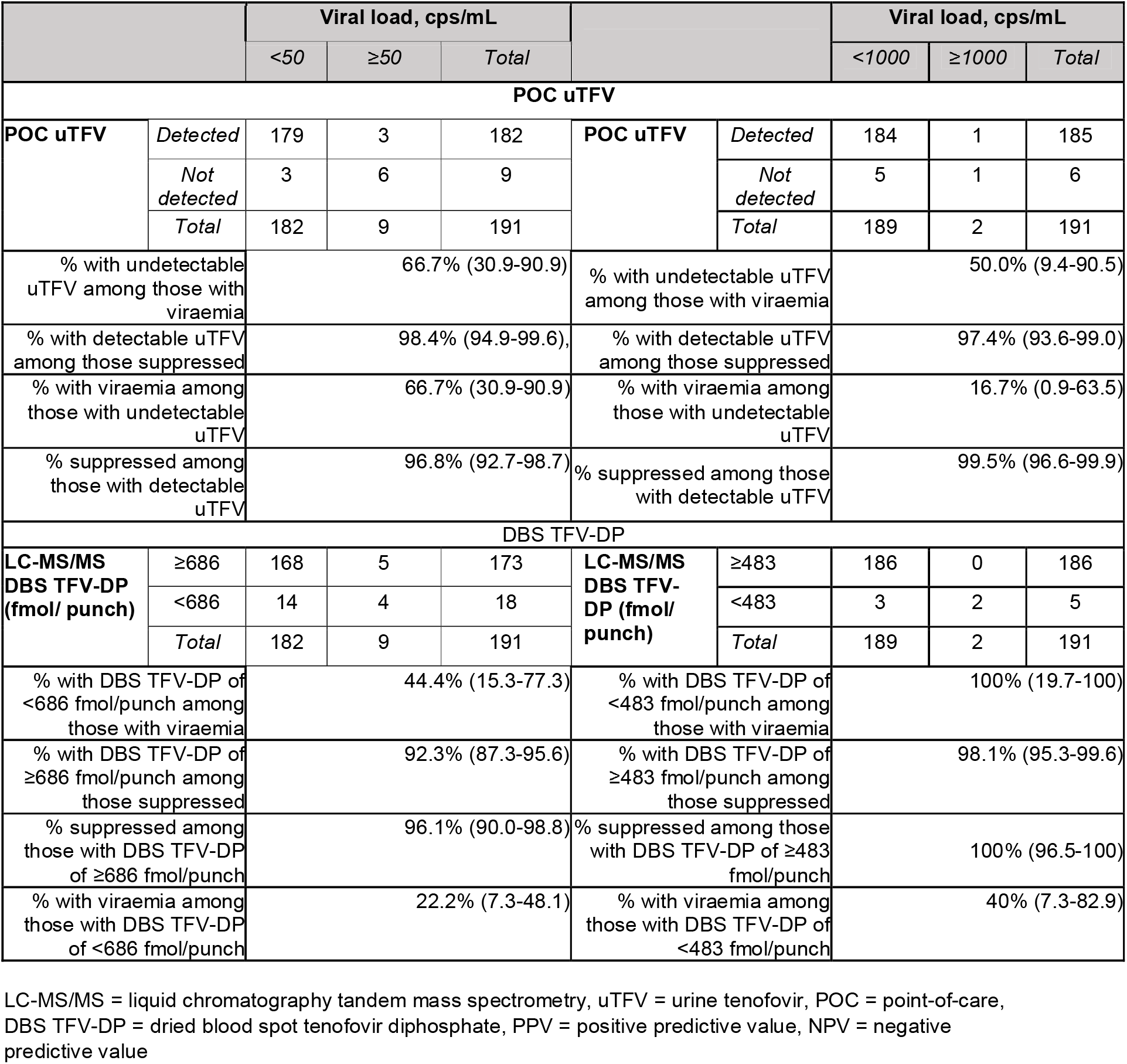
ASSOCIATION OF POINT-OF-CARE (POC) URINE TENOFOVIR (UTFV) AND DRIED BLOOD SPOT TENOFOVIR DIPHOSPHATE (DBS TFV-DP) CONCENTRATIONS WITH VIRAEMIA.

Results at VL ≥50 copies/mL were broadly similar to 1000 copies/mL, except that the proportion of those with viraemia who had low TFV-DP <686 fmol/punch was 44.4% (95% CI 15.3-77.3) (Table 2). Regarding retention-in-care, point-of-care uTFV (OR = 0.56, 95% CI 0.03-10.3) (Table S3) and DBS TFV-DP (OR = 1.02, 95% CI 0.95-1.08) were not associated with retention-in-care.

### Post-hoc analyses of association between point-of-care uTFV and DBS TFV-DP

Median (IQR) TFV-DP levels were substantially higher among samples with a point-of-care TFV detected result at 1295 fmol/sample (1005-1626 fmol/sample) compared to those with point-of-care TFV not detected result at 383 fmol/sample (89.8-551 fmol/sample) (p < 0.001) (Figure S3).

## Discussion

The point-of-care uTFV assay was accurate for detecting uTFV concentrations at the manufacturer’s threshold of 1,500 ng/mL. When applied to clinical outcomes in a community ART programme, while few people had viraemia both the point-of-care uTFV assay and DBS TFV-DP thresholds were associated with viral loads and identified >95% of those with viral suppression <1,000 copies/mL as ‘adherent’. Point-of-care uTFV results were not associated with retention in care by 16 weeks. Post-hoc analyses confirmed that detectable uTFV was associated with higher DBS TFV-DP concentrations.

Our findings are consistent with the two previous studies that assessed the diagnostic accuracy of the point-of-care uTFV assay. The first study, conducted as part of the assay development and validation [8], reported a sensitivity of 99% (95% CI 94-100) and specificity of 97% (95% CI 93-99) compared to LC-MS/MS. The second study, which we conducted in the POwER trial, reported a sensitivity of 96.1% (95% CI 90.0-98.8) and specificity of 95.2% (95% CI 75.3-100.0) [9]. While our study demonstrated a similarly high sensitivity of 99.5% (95% CI 96.5-100), our observed specificity was lower at 76.9% (95% CI 46.0-93.8), albeit with wide confidence intervals due to small numbers with low uTFV concentrations. A retrospective review of discordant test result images suggest that very faint lines on the lateral flow assay may have been missed, meaning results would have been interpreted as uTFV detected. Careful interpretation of results is therefore required, and, if necessary, expert review of borderline results.

Several other studies have also demonstrated an association between point-of-care urine tenofovir results and concurrent viraemia [9-12]. In our cohort, consisting of people previously found to be stable on ART and so eligible for a community ART delivery programme, viraemia was rare, resulting in very wide confidence intervals for our estimates of the proportion with viraemia who were identified as ‘non-adherent’ with undetectable point-of-care uTFV or low TFV-DP. However, the proportion with viral suppression with detectable uTFV/ high TFV-DP remained high, consistent with previous studies, reflecting that detectable urine tenofovir or high TFV-DP is strongly associated with viral suppression [9-12]. Other studies have shown that DBS TFV-DP concentrations are associated with concurrent [13, 14] and future VLs [6, 15]. Interestingly, we did not find associations between either TFV measure and future retention-in-care, suggesting that measures of adherence may be more useful for predicting future virologic outcomes, rather than engagement in care.

A major strength of this study is that it represents the first evaluation of point⍰of⍰care uTFV testing among people in community ART delivery programmes in South Africa, a setting where point-of-care diagnostics may be particularly useful. However, outcomes in this programme were generally good, with few participants experiencing viraemia or loss to follow⍰up. Another limitation is that tests were conducted in a laboratory setting rather than directly in the field, but this allowed better assessment of ideal performance diagnostic accuracy. Further evaluation in diverse settings is recommended.

Point⍰of⍰care uTFV testing offers a feasible option for more frequent adherence monitoring compared to routine VL testing, and could be integrated into community ART programmes to provide immediate feedback, target adherence interventions and triage people with undetectable uTFV for viral load monitoring [5]. While DBS TFV⍰DP concentrations measure longer-term adherence, the need for more resource⍰intensive LCMS currently limits applicability in community settings.

Our study demonstrates that the point⍰of⍰care uTFV assay is accurate, and that in community ART delivery programmes, both POC uTFV and DBS TFV-DP are highly specific for viral suppression. While these tests do not predict future retention in care, they may enhance programme monitoring by identifying individuals at risk of viraemia.

## Data Availability

All data produced in the present study are available upon reasonable request to the authors

## Acknowledgements

The authors would like to thank all participants in the study and acknowledge the work and support of staff at the Prince Cyril Zulu Clinic, eThekwini Municipality, CAPRISA and the National Health Laboratory Services at Addington and Inkosi Albert Luthuli Hospitals.

## References

1. Grimsrud A, Bygrave H, Doherty M, Ehrenkranz P, Ellman T, Ferris R, et al. Reimagining HIV service delivery: the role of differentiated care from prevention to suppression. Journal of the International AIDS Society. 2016;19(1):21484–

2. Dorward J, Drain PK, Garrett N. Point-of-care viral load testing and differentiated HIV care. Lancet HIV. 2018;5(1):e8–e9

3. Gandhi M, Bacchetti P, Rodrigues WC, Spinelli M, Koss CA, Drain PK, et al. Development and Validation of an Immunoassay for Tenofovir in Urine as a Real-Time Metric of Antiretroviral Adherence. EClinicalMedicine. 2018;2-3:22–8

4. Anderson PL, Liu AY, Castillo-Mancilla JR, Gardner EM, Seifert SM, McHugh C, et al. Intracellular Tenofovir-Diphosphate and Emtricitabine-Triphosphate in Dried Blood Spots following Directly Observed Therapy. Antimicrob Agents Chemother. 2018;62(1):1–13

5. Dorward J, Tlhaku K, Sookrajh Y, Munatsi P, Naidoo J, Tselana E, et al. Point-of-care HIV viral load testing in a community antiretroviral therapy delivery programme: A randomised controlled trial (PHILA). PLOS Global Public Health. 2026;6(3):e0005890

6. Dorward J, Govender K, Moodley P, Lessells R, Samsunder N, Sookrajh Y, et al. Urine tenofovir and dried blood spot tenofovir diphosphate concentrations and viraemia in people taking efavirenz and dolutegravir-based antiretroviral therapy. AIDS. 2024;38(5):697–702

7. Haldane JBS. The estimation and significance of the logarithm of a ratio of frequencies. Annals of Human Genetics. 1956;20(4):309–11

8. Gandhi M, Wang G, King R, Rodrigues WC, Vincent M, Glidden DV, et al. Development and validation of the first point-of-care assay to objectively monitor adherence to HIV treatment and prevention in real-time in routine settings. AIDS. 2020;34(2):255–60

9. Dorward J, Lessells R, Govender K, Moodley P, Samsunder N, Sookrajh Y, et al. Diagnostic accuracy of a point‐of‐care urine tenofovir assay, and associations with HIV viraemia and drug resistance among people receiving dolutegravir and efavirenz‐based antiretroviral therapy. Journal of the International AIDS Society. 2023;26(9):e26172

10. Marryshow TA, Muhairwe J, Tang A, Molulela MM, Matta R, Jordan MR. Determining the acceptability of point-of-care urine tenofovir testing and its performance in predicting HIV RNA suppression. International Journal of STD & AIDS. 2022;33(8):777–83

11. Hermans LE, Umunnakwe CN, Lalla-Edward ST, Hebel SK, Tempelman HA, Nijhuis M, et al. Point-of-care tenofovir urine testing for the prediction of treatment failure and drug resistance during initial treatment for human immunodeficiency virus type 1 (HIV-1) infection. Clinical Infectious Diseases. 2023;76(3):e553–e60

12. Van Zyl G, Jennings L, Kellermann T, Nkantsu Z, Cogill D, van Schalkwyk M, et al. Urine tenofovir-monitoring predicts HIV viremia in patients treated with high genetic-barrier regimens. AIDS. 2022;36(14):2057–62

13. Castillo-Mancilla JR, Morrow M, Coyle RP, Coleman SS, Gardner EM, Zheng JH, et al. Tenofovir diphosphate in dried blood spots is strongly associated with viral suppression in individuals with human immunodeficiency virus infections. Clinical Infectious Diseases. 2019;68(8):1335–42

14. Phillips TK, Myer L, Johnson LF, Jao J, Tiffin N, Orrell C, et al. A Comparison of Plasma Efavirenz and Tenofovir, Dried Blood Spot Tenofovir-Diphosphate, and Self-Reported Adherence to Predict Virologic Suppression Among South African Women. Journal of Acquired Immune Deficiency Syndromes. 2019;81(3):311–8

15. Jennings L, Robbins RN, Nguyen N, Ferraris C, Leu CS, Dolezal C, et al. Tenofovir diphosphate in dried blood spots predicts future viremia in persons with HIV taking antiretroviral therapy in South Africa. AIDS. 2022;36(7):933–40

