## Supplement for "Diagnostic accuracy of point-of-care urine tenofovir test, and associations between metrics of tenofovir use and treatment outcomes in a community ART programme"

**SUPPLEMENTARY FILE**

**Contents**

1. Liquid chromatography and dual tandem mass spectrometry (LC-MS/MS) sample analysis protocol
2. Table S1: Baseline characteristics of PHILA study participants, N = 196
3. Table S2: Discrepant point-of-care urine tenofovir test results
4. Table S3: Association of point-of-care urine tenofovir (POC TFV) with retention in care by 16 weeks
5. Figure S1: PHILA study CONSORT diagram
6. Figure S2: Point-of-care urine tenofovir (uTFV) tests of discrepant results
7. Figure S3: Box and whisker plot showing association between point-of-care uTFV result and DBS TFV-DP

### 1. Liquid chromatography and dual tandem mass spectrometry (LC-MS/MS) sample analysis protocol

We conducted LC-MS/MS at the Africa Health Research Institute in Durban, South Africa. A quantitative LC-MS/MS method was developed for the determination of tenofovir (TFV) in urine samples and TFV and tenofovir-diphosphate (TFV-DP) concentrations in dry blood spot (DBS) samples. The LC-MS/MS method was accurate, robust and quantitative over the concentration ranges; 0.5 - 80 µg/mL for TFV in urine and 200 - 8000 pg/mL for TFV and TFV-DP in DBS samples.

The urine and DBS samples were processed using a protein precipitation method. A 70% methanol:water (v/v) solution, which contained the deuterated internal standards; ^13^C labeled internal reference standards, ^13^C_5_-TFV and ^13^C_5_-TFV-DP was used for drug analyte extraction. The calibration standards and quality control solutions (containing TFV and TFV-DP) were prepared using the extraction solution.

The LC-MS/MS analysis was performed using an Agilent high pressure liquid chromatography (HPLC) system coupled to an AB Sciex 5500, triple quadrupole mass spectrometer equipped with an electrospray ionization (ESI) TurboIonSpray source. Analyst software, version 1.6.2 was used for data acquisition and quantitative data analysis.

Dry blood spot samples were analysed using ion pair-hydrophilic interaction chromatography coupled to tandem mass spectrometry (IP-HILIC-MS/MS) for the quantification of TFV and TFV-DP. The chromatographic separation was performed at a flow rate of 0.2 mL/min on a Luna Amino (NH2) column (Phenomenex, Torrance, CA) 100 mm × 2.0 mm, packed with 3.0 µm particles. Mobile phase A consisted of 100 mM hexafluoro-2-propanol (HFIP) and 0.5% diethylamine (DEA) (v/v) in water, and mobile phase B consisted of 0.1 M HFIP and 0.5% DEA (v/v) in acetonitrile. A sample volume of 5.0 µL was injected onto the HPLC column and the analytes were separated using a gradient elution. The autosampler syringe and the injection valve were washed with a water:acetonitrile (30:70, v/v) solution, post sample injection, to reduce carryover. The system was operated in negative-ion multiple reaction monitoring (MRM) mode set to detect precursor [M+H]^+^→ product ion transitions for TFV1 (*m/z* 285.8 → *m/z* 133.9), TFV2 (*m/z* 285.8→ *m/z* 151.0), TFV-DP1 (*m/z* 445.8 → *m/z* 158.9), TFV-DP2(*m/z* 445.8→ *m/z* 176.7) and the internal standard; ^13^C_5_-TFV *(m/z 290.9→m/z 139.0)* and ^13^C_5_-TFV-DP *(m/z 450.9→m/z 158.9)*. The optimized ESI source dependent parameters were set as follows; ion spray voltage (ISV): 5500V, temperature (TEM): 350°C, gas 1 (N_2_) and gas 2 (N_2_): 40 psi.

This LC-MS/MS assay is developed and validated according to US-FDA and ICH guidelines for bioanalytical assays. All reference drug standards and solvents are of high purity and LC-MS grade. Every analytical run includes calibration standards, quality control and system suitability samples and all samples are spiked with ^13^C stable isotope labeled internal reference standards for TFV and TFV-DP to ensure quantitative accuracy and precision. We have also successfully performed an inter-lab assay validation (with UCT Pharmacology) for the DBS assay.

2. **Table S1:** Baseline characteristics of PHILA study participants, N = 196

| **Variable** | **Levels** | **Total** |
| --- | --- | --- |
| Age, years | Median (IQR) | 44.0 (38.0 to 49.0) |
| Gender | Female, N(%) | 127 (64.8) |
|  | Male, N(%) | 69 (35.2) |
| Ethnicity | Black African, N(%) | 195 (99.5) |
|  | Other, N(%) | 1 (0.5) |
| Time since HIV diagnosis, years | Median (IQR) | 9.1 (6.7 to 12.0) |
| Time since ART initiation, years | Median (IQR) | 8.0 (6.0 to 10.9) |
| Initiation CD4 count, cells/µL | Median (IQR) | 249.5 (158.8 to 387.2) |
| Initiation CD4 count category, cells/µL | <200, N(%) | 62 (31.5) |
|  | 200-349, N(%) | 56 (28.6) |
|  | 350-499, N(%) | 31 (15.8) |
|  | >=500, N(%) | 23 (11.7) |
|  | (Missing), N(%) | 24 (12.2) |
| Current ART regimen at enrolment | TDF / 3TC / DTG, N(%) | 162 (82.7) |
|  | TDF / FTC / EFV, N(%) | 29 (14.8) |
|  | Other *, N(%) | 5 (2.6) |
| Time on current regimen, years | Median (IQR) | 2.7 (1.8 to 2.9) |
| POC TFV | Present, N(%) | 185 (94.4) |
|  | Absent, N(%) | 11 (5.6) |
| Urine tenofovir (uTFV), ng/mL | Median (IQR) | 21188(12335 to 40585) |
| Dried blood spot tenofovir diphosphate (DBS TFV-DP), fmol/punch | Median (IQR) | 1263.9 (955.6 to 1606.1) |
| DBS TFV-DP ≥ 483 | N(%) | 186 (97.4) |
| DBS TFV-DP ≥ 686 | N(%) | 168 (88) |
| Enrolment viral load, copies/mL | < 50, N(%) | 182 (92.9) |
|  | 50 - 999, N(%) | 5 (2.6) |
|  | >= 1000, N(%) | 2(1) |
|  | Missing, N(%) | 7 (3.6) |
| Retention by 16 weeks | Retained, N (%) | 169 (88) |

**5 adults with other regimens including AZT/3TC/EFV, ABC/3TC/DTG and AZT/3TC/LPVr*

**3. Table S2: Discrepant point-of-care urine tenofovir test results**

| **Discrepancy Number** | **Tenofovir Concentration (ng/mL)** | **Point-of-care urine tenofovir result** | **TFV-DP (fmol/punch)** |
| --- | --- | --- | --- |
| 1 | 641 | Detected | 444.4 |
| 2 | 912 | Detected | 995.1 |
| 3 | 1320 | Detected | 1803.3 |
| 4 | 2040 | Not detected | 1362.2 |

**4. Table S3: Association of point-of-care urine tenofovir (POC TFV) with retention in care by 16 weeks**

|  | | **Retention after 16 weeks** | | | |
| --- | --- | --- | --- | --- | --- |
|  |  | *Retained* | | *Not Retained* | *Total* |
| **POC TFV** | TFV not detected | 6 | | 0 | 6 |
|  | TFV detected | 163 | | 22 | 185 |
|  | *Total* | 169 | | 22 | 191 |
|  | Odds ratio | | 0.56 (0.03-10.3) | | |

5. Figure S1: PHILA study CONSORT diagram

POC = point-of-care; uTFV= Urine tenofovir; LCMS= Liquid chromatography and dual tandem mass spectrometry; TFV-DP=Tenofovir Diphosphate

200 adults randomized in PHILA

- (n = 200)

Excluded (n=4)

• No POC uTFV result

196 adults included in POC uTFV diagnostic accuracy analysis

191 adults included in POC uTFV and DBS TFV-DP association with viraemia and retention-in-care analysis

Excluded (n=5)

• Not on TDF-containing ART

**6. Figure S2: Point-of-care urine tenofovir (uTFV) tests of discrepant results**

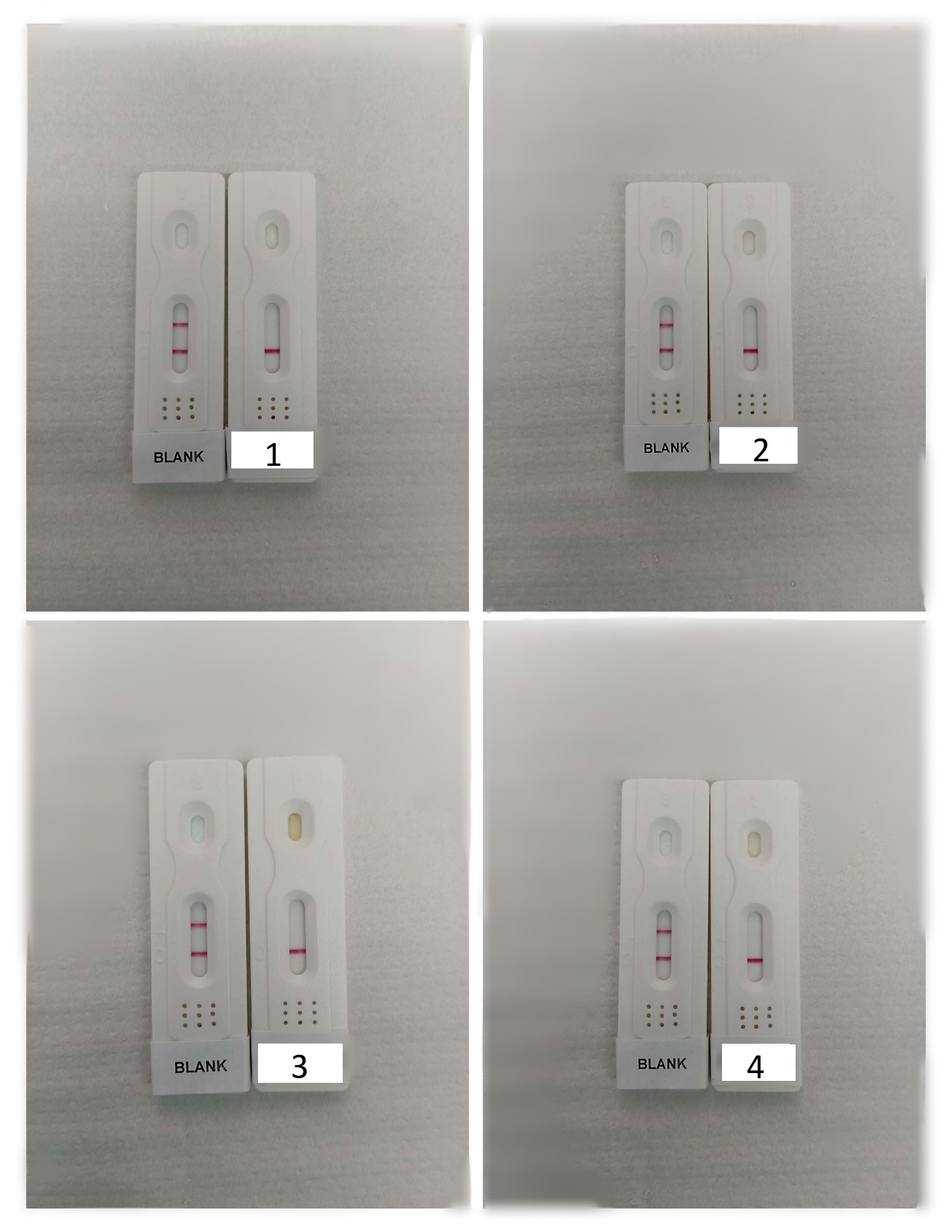

**7. Figure S3: Box and whisker plot showing association between point-of-care tenofovir (TFV) result and tenofovir diphosphate (TFV-DP)**

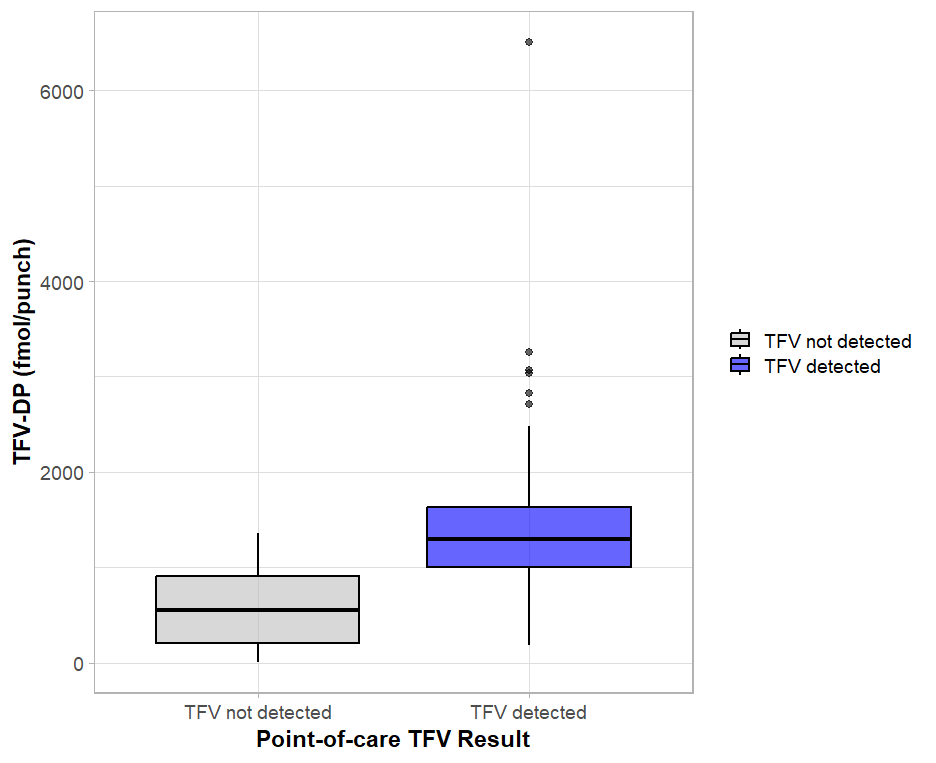
